# PREVALENCE OF MOLECULAR AND SEROLOGICAL TESTS OF THE NEW CORONAVIRUS (SARS-CoV-2) IN CARLOS CHAGAS-SABIN LABORATORIES IN CUIABÁ

**DOI:** 10.1101/2020.10.26.20219683

**Authors:** Cristiane Coimbra de Paula, João Pedro Castoldo Passos, Walkiria Shimoya-Bittencour, Caroline Aquino Vieira de Lamare, Ruberlei Godinho de Oliveira

## Abstract

**BACKGROUND:** COVID-19, the disease caused by the new coronavirus (SARS-CoV-2), became a pandemic in 2020 with mortality rate of 2% and high transmissibility, which makes studies with an epidemiological profile essential.

**OBJECTIVES:** To characterize the population that performed the SARS-CoV-2 molecular and serological tests in Carlos Chagas-Sabin laboratories in Cuiabá.

**METHODS:** A retrospective cross-sectional study was carried out with all the samples collected from nasal swab tested by RT-PCR and serological for SARM-CoV-2 IgM / IgG from the population served between April and July 2020.

**FINDINGS:** In the analyzed period, 11,113 PCR-Covid-19 exams were registered. Of this total of cases, 3,912 (35.20%) tested positive, while 6,889 (61.90%) did not detect viral RNA and 312 (2.80%) of the visits resulted as undetermined. The peak of positive RT-PCR performed in July (n = 5878), with 35.65% (n = 2096). A total of 6,392 tests performed on SOROVID-19, with a peak of 1161 (18.16%) of the positive tests for SARS-CoV-2 in July.

**MAIN CONCLUSIONS:** Molecular positivity and serological tests, both peaked in July 2020, were mostly present in women aged 20-39, characterizing Cuiabá as the epicenter of the Midwest region in this period due to the high rate of transmissibility of SARS-CoV-2.

## Introduction

COVID-19 the disease caused by the new coronavirus (SARS-CoV-2) became an alarming threat to public health in 2020, despite global efforts to prevent its spread.^1^ SARS-CoV-2 spread rapidly, reaching more than 100 countries in five continents, forcing the World Health Organization (WHO) to declare COVID-19 as a pandemic on March 11, 2020 ^2,3^.

This virus has an RNA virus of the order Nidovirales of the family Coronaviridae that can induce disease in the upper, and eventually, lower respiratory tract in vulnerable patients, immunocompromised patients with chronic diseases, the elderly and, sporadically, in children and young people ^4^. After this contact, there is an average infection incubation period of 5 days, with an interval of up to 12 days. There are other reports that indicate an interval of 7 to 14 days, but not very precise so far, once the viral spreading happens directly among humans through community transmission ^5^. COVID-19 is a new disease that deserves special attention and care because the symptoms among infected people, from mild to severe, with mortality estimated at just over 2% 3. However, in Várzea Grande, a city located in the Baixada Cuiabana, there was a high lethality period (6-10%) from April to July 2020 according to the data released by the Municipal Health Department. The State Department of Health (SES-MT) has notified, to date, 83,490 confirmed cases in Cuiabá, Várzea Grande and thirteen more cities in Mato Grosso, that are classified as “very high” risk for SARS-CoV-2 and with a total of 2614 deaths so far. The number of new cases registered in the first week of August was 42% higher than the same rate in the first week of July, classifying the state as the epicenter of COVID-19 cases. (http://www.saude.mt.gov.br/painelcovidmt/).

The transmission of this virus occurs quickly through aerosols in patients undergoing airway procedures, such as orotracheal intubation or airway aspiration. Direct contact with mucous membranes (mouth, nose or eyes) contributes to the invasion and pathogenicity of SARS-CoV-2. Thus, some population groups are more vulnerable to being affected by the disease, due to general conditions, ranging from health conditions to the way of life to which they are exposed^6^. Patients who meet the criteria for suspected cases should be tested for SARS-CoV-2, using samples collected from the nasopharyngeal mucous by the nasal swab ^6,7^. The SARS-CoV-2 RNA is detected by reverse transcription polymerase chain reaction (RT-PCR) and a positive SARS-CoV-2 test confirms the diagnosis of COVID-19 ^8^.

However, if the initial test is negative, but suspected of COVID-19, WHO recommends resampling and testing of various airway locations, as well as testing for antibodies ^9^. Serological tests are used to assess the window period in the stage of current (IgM) or chronic (IgG) infection.

The tests are based on the principle of lateral flow immunoassay for the detection of IgG / IgM antibodies against SARS-CoV-2 in whole blood, serum and plasma of humans, requiring quantification according to the onset of symptoms reported by the patient to avoid false negative results^10^. Thus, identifying the magnitude of the health problem in the population is the first step towards the development of effective decision-making strategies in evidence-based public health situations^11, 12^, as well as understanding the spatial distribution of the disease is fundamental for the development of strategies during the early stages of the COVID-19 ^13^ emergency.

In this regard, among the clinical and laboratory repercussions of the patient with COVID-19, this study aims to characterize the prevalence of molecular (RT-PCR) and serological (IgM and IgG) tests for SARS-CoV-2 performed in Carlos Chagas-Sabin laboratories in Cuiabá from April to July 2020.

## Materials and methods

This is a retrospective cross-sectional study with samples from the secondary database of Carlos Chagas Laboratory - Grupo Sabin, in Cuiabá - MT, collected between April and July 2020. In order to verify the prevalence of molecular and serological tests for SARS-CoV-2, all samples from people seen in the laboratory of both sexes were included, without age restriction; and the reports of the molecular tests performed by qPCR-RT through the extraction of the genetic material of the nasal swab virus, as well as serological tests SOROVID-19 (IgM and IgG) for the detection of antibodies to SARS-CoV-2. Rapid IgG or IgM test data were excluded from the study. Data were expressed as absolute frequency and percentages with tabulation in Microsoft Excel.

This study was approved by the Ethics Committee of UNIVAG-Centro Universitário under protocol number CAAE: 37320320.1.0000.5692. The procedures followed were in accordance with the ethical standards of the responsible committee on human experimentation (institutional or regional). It is the Principal Researcher’s responsibility to ensure that all researchers associated with this project are aware of the conditions of approval and which documents have been approved.

## Results

Between April and July 2020, 11,113 PCR-Covid-19 tests were registered in Carlos Chagas-Sabin laboratories located in Cuiabá. Out of the total number of cases, 3,912 (35.20%) tested positive, while 6,889 (61.90%) did not have the disease and 312 (2.80%) of the visits resulted in indeterminate (Table 1).

**Table 1.** Prevalence of COVID-19 tests performed in Carlos Chagas-Sabin laboratories in Cuiabá from April to July 2020.

| Variables | N = 11.113 | % |
| --- | --- | --- |
| Detected | 3.912 | 35.20 |
| Not detected | 6.889 | 61.99 |
| Undetermined | 312 | 2.80 |

Subtitle: %: Percentage

In the same period, 6,392 SOROVID-19 tests (IgG and IgM) were recorded at the Carlos Chagas-Sabin laboratory in the municipality of Cuiabá. Of the total number of cases, 1,413 (22.10%) tested positive for both testicles and / or only for IgG or IgM, with IgG late in contact with the disease, immunity against viruses and IgM recent contact with viruses that the sick person transmits when in contact with other people who live together, while 4,979 (77.89%) of the tests were non-reactive (Table 2).

**Table 2.** Prevalence of COVID-19 tests performed in Carlos Chagas-Sabin laboratories in Cuiabá from April to July de 2020.

| Variables | N = 6.392 | % |
| --- | --- | --- |
| Reactive | 1.413 | 22.10 |
| Non-reactive | 4.979 | 77.89 |

Subtitle: %: Percentage

There was a predominance of cases in 6474 women (58.2%) in the 20-49-year-old age group. Positive cases registered with a peak in July (n = 2096), totaling 5680 exams performed in that month for COVID-19th, undetected (n = 3584) and undetermined (n = 198) (Table 3).

**Table 3.**
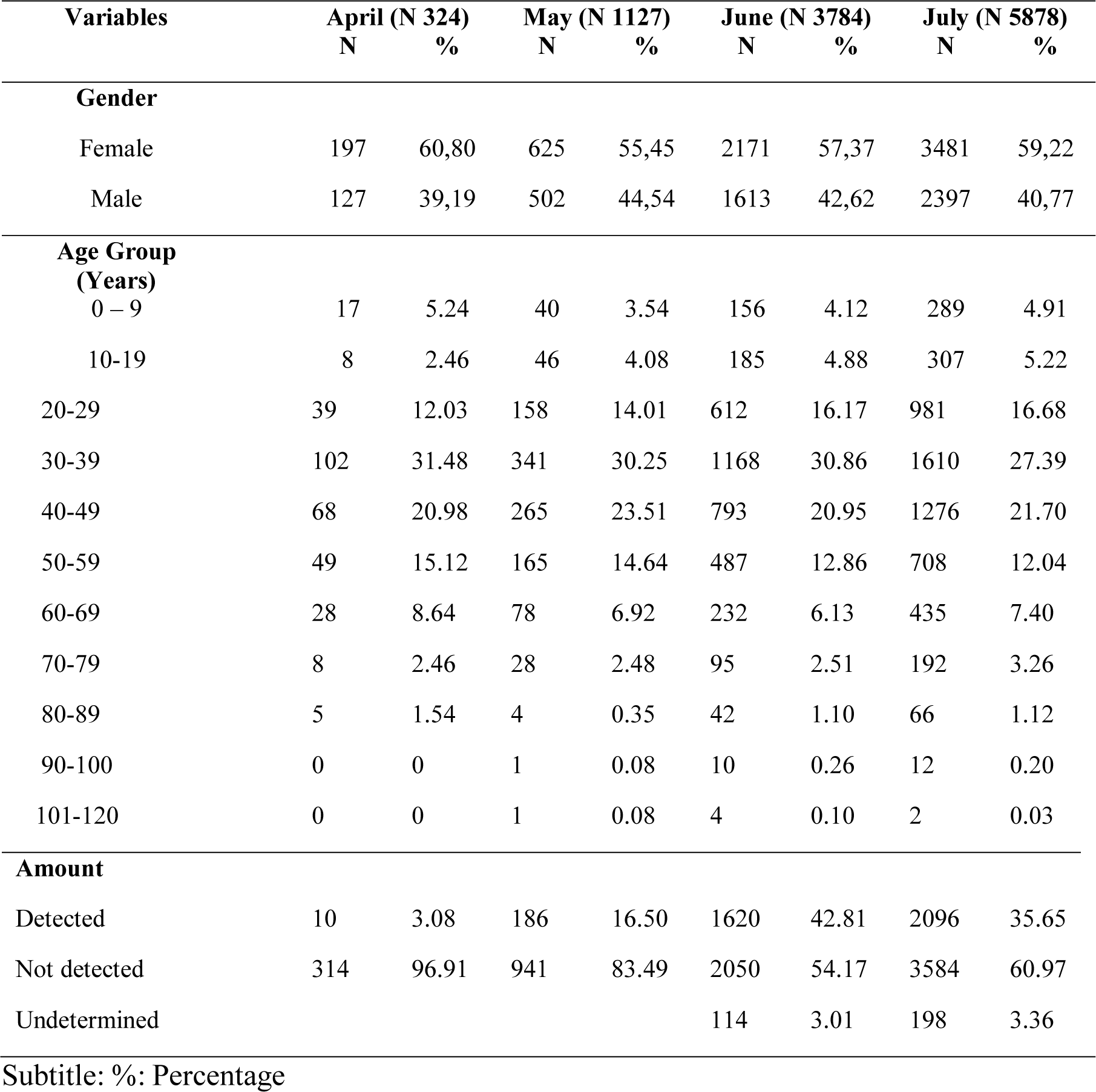
Prevalence of cases of COVID-19, disease caused by the new coronavirus, (SARS-CoV-2) performed in Carlos Chagas-Sabin Laboratories in Cuiabá per month, gender and age group.

In April, 324 tests were registered, 10 (3.08%) of which were confirmed by COVID-19 and 314 (96.91%) were negative for the disease. Of the total cases, 197 (60.80%) were female and 127 (39.19%) were male, individuals between 20 and 59 years of age were the ones who most performed the exams (Table 3).

In May, 1127 tests were recorded, 186 (16.50%) of which were confirmed COVID-19 and 941 (83.49%) were negative for the disease. Of the total number of cases, 625 (55.45%) were female and 502 (44.54%) were male, individuals between 20 and 59 years of age were the ones who most performed the exams in that month (Table 3). In June, 3784 tests were registered, of which 1620 (42.81%) were confirmed COVID-19 and 2050 (54.17%) were negative for the disease and 114 (3.01%) had undetermined results, those ones being believed to have been performed outside the appropriate period for the exam. Of the total number of cases, 2171 (57.35%) were female and 1613 (42.62%) were male, aged between 20 and 59, being the ones who most performed the exams in that month (Table 3).

In July, 5878 tests were registered, 2096 (35.65%) were confirmed positive for COVID-19 and 3584 (60.97%) were negative for the disease and 198 (3.36%) had undetermined results, those ones being believed to have been performed outside the appropriate period for the exam. Of the total cases, 3481 (59.22%) were female and 2397 (40.77%) were male, remaining the same range of individuals aged 20 to 59 being the ones looking for the laboratories to perform the exams (Table 3).

The total of 6,392 tests performed by SOROVID19 at the Carlos Chagas-Sabin Laboratory experienced a significant increase in the total number of tests collected per month, subsequent to April, due to the high rate of transmission and community circulation of SARS-CoV-2 (Table 4). In April, 202 tests were recorded, 14 (6.93%) were confirmed for SOROVID and 188 (93.06%) were negative for the disease. Of the total cases, 110 (54.45%) were female and 92 (45.54%) were male, individuals between 30 and 59 years of age were the ones who most performed the exams. In May, 496 tests were recorded, 23 of which (4.63%) were confirmed for SOROVID and 473 (95.36%) were negative for the disease. Of the total number of cases, 296 (59.67%) were female and 200 (40.32%) were male, individuals between 20 and 59 years of age were the ones who performed the exams in that month. In June, 1,446 tests were recorded, 215 (14.86%) were confirmed for SOROVID and 1231 (85.13%) were negative for the disease (Table 4).

**Table 4.** Prevalence of cases of COVID-19, disease caused by the new coronavirus, (SARS-CoV-2) performed in Carlos Chagas-Sabin Laboratories in Cuiabá per month, gender and age group.

| Variables | April (N 202) |  | May (N 496) |  | June (N 1446) |  | July (N 4248) |  |
| --- | --- | --- | --- | --- | --- | --- | --- | --- |
|  | N | % | N | % | N | % | N | % |
| **Gender** |  |  |  |  |  |  |  |  |
| Female | 110 | 54.45 | 296 | 59.67 | 868 | 60.02 | 2545 | 59.83 |
| Male | 92 | 45.54 | 200 | 40.32 | 578 | 39.97 | 1706 | 40.16 |
| **Age Group** |  |  |  |  |  |  |  |  |
| **(Years)** |  |  |  |  |  |  |  |  |
| 0 – 9 | 8 | 3.96 | 14 | 2.82 | 48 | 3.31 | 98 | 2.30 |
| 10-19 | 8 | 3.96 | 26 | 5.24 | 56 | 3.87 | 130 | 3.06 |
| 20-29 | 14 | 6.93 | 56 | 11.29 | 146 | 10.09 | 566 | 13.32 |
| 30-39 | 58 | 28.71 | 142 | 28.62 | 404 | 27.93 | 958 | 22.55 |
| 40-49 | 44 | 21.78 | 130 | 26.20 | 356 | 24.61 | 942 | 22.17 |
| 50-59 | 32 | 15.84 | 60 | 12.09 | 212 | 14.66 | 772 | 18.17 |
| 60-69 | 30 | 14.85 | 54 | 10.88 | 158 | 10.92 | 506 | 11.91 |
| 70-79 | 8 | 3.96 | 12 | 2.41 | 50 | 3.45 | 202 | 4.75 |
| 80-89 | 0 | 0 | 2 | 0.40 | 16 | 1.10 | 56 | 1.31 |
| 90-100 | 0 | 0 | 0 | 0 | 0 | 0 | 14 | 0.32 |
| 101-120 | 0 | 0 | 0 | 0 | 0 | 0 | 4 | 0.09 |
| **Amount** |  |  |  |  |  |  |  |  |
| Reactive | 14 | 6.93 | 23 | 4.63 | 215 | 14.86 | 1161 | 27.33 |
| Non-reactive | 188 | 93.06 | 473 | 95.36 | 1231 | 85.13 | 3087 | 72.66 |

Subtitle: %: Percentage.

Of the total number of cases, 868 (60.02%) were female and 578 (39.97%) were male, individuals between 20 and 59 years of age were the ones who most performed the exams in that month. In the results of July, 4248 tests were recorded, 1161 (27.33%) were confirmed positive for SOROVID and 3087 (72.60%) negative for the disease. Of the total cases, 2545 (59.83%) were female and 1706 (40.16%) were male, remaining in the same range of individuals aged 20 to 59 years (Table 4).

## Discussion

The results of the 11,113 tests performed for SARS-CoV-2 in the period from April to July, detected 35.20% positive by the molecular test (RT-PCR) and 22.10% of the serological tests by the SOROVID-19 test (IgG / IgM) peaked in July 2020, being the women the majority in the 20-39 age group. Those data demonstrate a progressive increase in cases, associated with the high rate of transmission and community circulation of SARS-CoV-2 in Cuiabá during the analyzed period, characterizing the month of July as the epicenter of COVID-19 in the central west region of Brazil ^14,15.^

Regarding gender, the incidence of SARS-CoV-2 was more positive in women, similar to the findings in another study conducted in Mato Grosso^19^ and Rio de Janeiro^10^ which also reported the majority of cases in women (51.4%), whereas in males it was 47.7%. In contrast to our study in Wuhan province, China^20^, the prevalence was higher in men (56%) with mortality 56.5% and 38.0% of female deaths^10^.

According to data from the State Department of Health, Cuiabá, Várzea Grande and 13 other cities in Mato Grosso are classified as “very high” risk for the new coronavirus. This risk classification points to cities with more than 150 active cases on that date, such as Sorriso with 24.81%, Barra do Garças with 19.92% and Paranatinga with 14.83%, the other cities in the State are between 2-11 %. In Cuiabá, there are 13,958 confirmed cases with 636 deaths and the city Várzea Grande with a high mortality rate, as there are 5,234 confirmed cases with 337 deaths and a 7% lethality rate, confirming that the Baixada Cuiabana has a lethality rate above the national average.

According to the Epidemiological Bulletin of the State of Mato Grosso, the profile of patients with COVID-19 is predominantly female (52%), as well as the prevalence of deaths is also higher in women (59.3%). However, this distribution differs between the states, as in Maranhão the predominance of deaths by COVID-19 was male (62%)^16^. It is believed that women seek health services more frequently than men, and there may be underreporting of cases in the male population, as, historically, men seek health services less, which can lead to the worsening of the disease, late treatment and evolution to death.

Regarding the age group, there was a predominance of cases of patients between 30 and 49 years old for both tests, molecular and serological, for the detection of SARS-CoV-2. Those findings are similar to the ones found in a study carried out in Maranhão (28.4%)^16^ and in Wenzhou (China), which presented 58.9% of cases in that same age group^17^. Likewise, individuals aged 30 to 59 years were more prevalent among the cases studied in Rio de Janeiro^10^. It is worth emphasizing the need to endorse non-pharmacological measures, in order to reduce the number of people with the disease in that age group, which characterizes the economically active population, reinforces the adoption of assertive socioeconomic measures and preventive measures with the epidemiological surveillance of each citizen in order to decrease the transmissibility of SARS-CoV-2 19.

The serological tests for the detection of antibodies were mostly non-reactive, probably due to sample collection during the immunological window period, as well as in asymptomatic patients or those who did not report the onset period of symptoms for SARS-CoV-2 IgM / IgG positivity as recommended by the Ministry of Health (10-12 days).

Among the limitations, despite the secondary data of this study being collected in a locally and nationally known laboratory, the samples are representative and descriptive only from the city of Cuiabá and region roundabout. This in fact precludes a statewide coverage of the epidemiology of COVID-19, as well as the possibility that the population may have performed tests for SARS-CoV-2 in other laboratories available in the capital. However, it is one of the first studies describing the cases of COVID-19 and the type of approach carried out in Cuiabá which directly contributes to decision-making by requiring notification to the surveillance and health control bodies.

## Conclusion

Therefore, we conclude that the prevalence of COVID-19 in Cuiabá - MT was higher in women, aged 30 to 39 years, and the number of confirmed cases was higher from June to July 2020. The amount of Detection of exams by RT-PCR and reagents for SOROVID (IgM and IgG) monthly increased having its peak in July 2020, which in fact reflects the high transmissibility rate of SARS-CoV-2 in Cuiabá with a public health emergency.

## Data Availability

This is a retrospective cross-sectional study with samples from the secondary database of Carlos Chagas Laboratory - Grupo Sabin, in Cuiab&aacute - MT, collected between April and July 2020. In order to verify the prevalence of molecular and serological tests for SARS-CoV-2, all samples from people seen in the laboratory of both sexes were included, without age restriction; and the reports of the molecular tests performed by qPCR-RT through the extraction of the genetic material of the nasal swab virus, as well as serological tests SOROVID-19 (IgM and IgG) for the detection of antibodies to SARS-CoV-2. Rapid IgG or IgM test data were excluded from the study. Data were expressed as absolute frequency and percentages with tabulation in Microsoft Excel.

## Acknowledgements

We thank Laboratório Carlos Chagas-Sabin in Cuiabá, MT, Brazil for technical support durgin the analysis and Ethics committee to approval the project.

## Author’s contribution

CCP, CAVL and RGO designed the study. JPCP and WSB performed the experiments. CC de P, WSB and RGO wrote the manuscript. CCP and RGO coordinated the study. All authors contributed to data discussion and in the final manuscript.

## References

1. PIZZICHINI, Marcia Margaret Menezes, PATINO, Cecilia Maria, FERREIRA, Juliana Carvalho. Medidas de frequência: calculando prevalência e incidência na era do COVID-19. J Bras Pneumol. 2020;46(3):243.

2. Roberto P, Stephens S. Capítulo 2 Virologia. Conceitos e Métodos para a Formação Profissionais em Laboratórios Saúde. 2010;125–220.

3. Zhu N, Zhang D, Wang W, et al. A novel coronavirus from patients with pneumonia in China, 2019. New England Journal of Medicine, 2020. doi 10.1056/NEJMoa2001017.

4. Cui J, Li F, Shi ZL. Origin and evolution of pathogenic coronaviruses. Nature Reviews Microbiology 2019; 17:181–92.

5. World Health Organization. Novel Coronavirus (2019-nCoV) technical guidance.

6. Patel A, Jernigan DB, 2019-nCoV CDC Response Team. Initial Public Health Response and Interim Clinical Guidance for the 2019 Novel Coronavirus Outbreak - United States, December 31, 2019-February 4, 2020. MMWR Morb Mortal Wkly Rep 2020; 69:140.

7. Interim Guidelines for Collecting, Handling, and Testing Clinical Specimens from Persons Under Investigation (PUIs) for Coronavirus Disease 2019 (COVID-19). February 14, 2020 https://www.cdc.gov/coronavirus/2019-nCoV/lab/guidelines-clinical-specimens.html (Acessado em 21 de Julho, 2020).

8. WHO. Coronavirus disease (COVID-19) technical guidance: Surveillance and case definitions. https://www.who.int/emergencies/diseases/novel-coronavirus-2019/technical-guidance/surveillance-and-case-definitions (Acessado em 21 de julho, 2020).

9. Hallal, Pedro Curi; Horta, Bernardo L; Barros, Aluísio J D; Dellagostin, Odir A; Hartwig, Pellanda, Fernando P. Lúcia C. Struchiner, CláUdio José; Burattini, Marcelo N; Silveira, Mariângela Freitas Da; Meneze, S Ana M B; Barros, Fernando C; Victora, Cesar Gomes. Evolução da prevalência de infecção por COVID-19 no Rio Grande do Sul, Brasil: inquéritos sorológicos seriados. Ciência & Saúde Coletiva, 25(Supl.1):2395–2401, 2020.

10. Cavalcante, João Roberto E Abreu, Ariane De Jesus Lopes de. COVID-19 no município do Rio de Janeiro: análise espacial da ocorrência dos primeiros casos e óbitos confirmados. Epidemiol. Serv. Saúde [online]. 2020, vol.29, n.3.

11. Chinazzi M, Davis JT, Ajelli M, Gioannini C, Litvinova M, Merler S, et al. The effect of travel restrictions on the spread of the 2019 novel coronavirus (COVID-19) outbreak. Science [Internet]. 2020 [acesso 2020 Ago 06]; 1-11. Disponível em: https://science.sciencemag.org/content/early/2020/03/05/science.aba9757

12. Ministério da Saúde (BR). Secretaria de Atenção Primária à Saúde - SAPS. Protocolo de manejo clínico do coronavírus (covid-19) na atenção primária à saúde [Internet]. Brasília: Ministério da Saúde; 2020 [citado 2020 mar 28]. 40 p. Disponível em: https://www.unasus.gov.br/especial/covid19/pdf/37

13. Guan W, Ni Z, Hu Y, Liang W, Ou C, He J, et al. Clinical characteristics of coronavirus disease 2019 in China. N Engl J Med [Internet]. 2020 Apr [cited 2020 May 6];382:1708-20. Available from: https://doi.org/10.1056/NEJMoa2002032

14. Ministério da Saúde (BR). Centro de Operações de Emergências em Saúde Pública | COE-COVID-19. Plano de contingência nacional para infecção humana pelo novo coronavírus COVID-19 [Internet]. Brasília: Ministério da Saúde; 2020 [citado 2020 abr 2]. 24 p. Disponível em: https://portalarquivos2.saude.gov.br/images/pdf/2020/agosto/11/plano-contingenciacoronavirus-COVID19.pdf10.

15. Croda JHR, Garcia LP. Respuesta inmediata de la vigilancia en salud a la epidemia de COVID-19. Epidemiol Serv Saúde [Internet]. 2020 agost [citado 2020 abr 6];29(1):e2020002. Disponível em: https://doi.org/10.5123/s1679-49742020000100021

16. Almeida, Joelson dos Santos; CARDOSO, Jonas Alves; Cordeiro, Eduardo Costa; Lemos, Messias; AraÚJo, Telma Maria Evangelista; Sardinha, Ana Hélia De Lima. CARACTERIZAÇÃO EPIDEMIOLÓGICA DOS CASOS DE COVID-19 NO MARANHÃO: UMA BREVE ANÁLISE. Revista Prevenção de Infecção e Saúde, maio 2020.

17. Yi Han, BS; Yi Liu, BS; Liyuan Zhou,Enguo Chen, Pengyuan Liu, Xiaoqing Pan, Yan Lu. Epidemiological Assessment of Imported Coronavirus Disease 2019 (COVID-19) Cases in the Most Affected City Outside of Hubei Province,Wenzhou, China JAMA Network Open. 2020; 3(4):e206785. doi:10.1001/jamanetworkopen.2020.6785

18. Rezer F, Faustino WR, Maia CS. Incidence of COVID-19 in the mesoregions of the state of Mato Grosso: confirmed and notified cases. Rev Pre Infec e Saúde [Internet]. 2020;6: 10317. Available from: http://www.ojs.ufpi.br/index.php/nupcis/article/view/10317 doi: https://doi.org/10.26694/repis.v6i0.10317 [In Press].

19. Instituto Brasileiro de Geografia e Estatística (IBGE). Pesquisa nacional por amostra de domicílios de 2001 a 2015. Microdados. Rio de Janeiro: IBGE. 2017

20. Li Q, Guan X, Wu P, Wang X, Zhou L et al. Early Transmission Dynamics in Wuhan, China, of Novel Coronavirus–Infected Pneumonia. N Engl J Med. 2020; 26 (382):1199–1207. DOI: 10.1056/NEJMoa 2001316.

