## Supplemental Data 1 for "PREVALENCE OF MOLECULAR AND SEROLOGICAL TESTS OF THE NEW CORONAVIRUS (SARS-CoV-2) IN CARLOS CHAGAS-SABIN LABORATORIES IN CUIABÁ"

### PARECER CONSUBSTANCIADO DO CEP

#### DADOS DO PROJETO DE PESQUISA

**Título da Pesquisa:** PREVALÊNCIA DOS CASOS DE COVID-19 (DOENÇA CAUSADA PELO CORONAVÍRUS SARS-COV-2) REALIZADOS NO LABORATÓRIO CARLOS CHAGAS ¿ GRUPO SABIN EM CUIABÁ NO PERÍODO DE FEVEREIRO A JULHO DE

**Pesquisador:** Cristiane Coimbra de Paula

**Área Temática:**

**Versão:** 2

**CAAE:** 37320320.1.0000.5692

**Instituição Proponente:** INSTITUICAO EDUCACIONAL MATOGROSSENSE-IEMAT

**Patrocinador Principal:** Financiamento Próprio

#### DADOS DO PARECER

**Número do Parecer:** 4.302.651

**Apresentação do Projeto:**

O projeto encontra-se bem organizado e bem escrito.

**Objetivo da Pesquisa:**

Os objetivos estão bem definidos.

**Avaliação dos Riscos e Benefícios:**

Por ser uma pesquisa retrospectiva, os riscos são mínimos quando comparados aos benefícios.

**Comentários e Considerações sobre a Pesquisa:**

A pesquisa possui grande relevância devido à importância que o tema assumiu no ano de 2020.

**Considerações sobre os Termos de apresentação obrigatória:**

As solicitações prévias foram atendidas.

**Recomendações:**

Aprovar.

**Conclusões ou Pendências e Lista de Inadequações:**

Sem pendências.

**Considerações Finais a critério do CEP:**

Aprovação recomendada pelo CEP.UNIVAG. A partir de agora o pesquisador poderá dar inicio a suas atividades, devendo nos enviar os relatórios parciais e finais conforme seu cronograma previsto. Cordialmente,CEP.UNIVAG

**Este parecer foi elaborado baseado nos documentos abaixo relacionados:**

| Tipo Documento | Arquivo | Postagem | Autor | Situação |
|---|---|---|---|---|
| Informações Básicas do Projeto | PB_INFORMAÇÕES_BÁSICAS_DO_PROJETO_1600145.pdf | 21/09/2020 19:16:02 | | Aceito |
| Declaração de Pesquisadores | declaracaoparticipacaodospesquisadoresprojetodepesquisa.pdf | 21/09/2020 19:14:47 | Cristiane Coimbra de Paula | Aceito |
| Declaração de Instituição e Infraestrutura | Declaracao_instituicao_numeradoerubrica.pdf | 21/09/2020 19:08:53 | Cristiane Coimbra de Paula | Aceito |
| Projeto Detalhado / Brochura Investigador | PROJETO_COVID19_CARLOSCHAGAS_RUBERLEI_COMITEETICA.docx | 21/09/2020 19:08:26 | Cristiane Coimbra de Paula | Aceito |
| Outros | formulariodeencaminhamentodoprojetoaoCEP_UNIVAG.docx | 28/07/2020 15:19:22 | Cristiane Coimbra de Paula | Aceito |
| Outros | Solicitacaodedispensa_TCLE.docx | 28/07/2020 15:18:59 | Cristiane Coimbra de Paula | Aceito |
| Outros | Termodeautorizacaoparautilizacaomanuseiodados.pdf | 28/07/2020 15:18:35 | Cristiane Coimbra de Paula | Aceito |
| Outros | CurriculoLattes_JoaoPedroCastoldoPassos.pdf | 27/07/2020 12:02:02 | Cristiane Coimbra de Paula | Aceito |
| Folha de Rosto | folhaDeRostoassinada.pdf | 27/07/2020 11:44:50 | Cristiane Coimbra de Paula | Aceito |
| Outros | CurriculoLattes_WalkiriaShimoya.pdf | 27/07/2020 11:41:25 | Cristiane Coimbra de Paula | Aceito |
| Outros | CurriculoLattes_RuberleiGodinhodeOliveira.pdf | 27/07/2020 11:40:56 | Cristiane Coimbra de Paula | Aceito |
| Outros | CurriculoLattes_CristianeCoimbradePaula.pdf | 27/07/2020 11:40:15 | Cristiane Coimbra de Paula | Aceito |

**Situação do Parecer:**
Aprovado

**Necessita Apreciação da CONEP:**
Não

**Endereço:** Av. Dom Orlando Chaves nº 2655
**Bairro:** CRISTO REI **CEP:** 78.118-000
**UF:** MT **Município:** VARZEA GRANDE

VARZEA GRANDE, 28 de Setembro de 2020

---

**Assinado por:**

**Rosa Maria Elias**
**(Coordenador(a))**

**Endereço:** Av. Dom Orlando Chaves nº 2655
**Bairro:** CRISTO REI **CEP:** 78.118-000
**UF:** MT **Município:** VARZEA GRANDE
